# Validation of “Days Alive and out of Hospital” as an Outcome Measure After Coronary Artery Bypass Graft Surgery, Acute Coronary Syndrome and Heart Failure Hospitalisation

**DOI:** 10.1101/2024.02.12.24302587

**Authors:** Robert Grant, Weiqi Liao, Joanne Miksza, Marius Roman, Gavin Murphy

## Abstract

**Background and Rationale:** ‘Days alive and out of hospital’ (DAOH) is a composite outcome measure that integrates several outcomes, including death, hospital length-of-stay, and hospital readmission. The minimum clinical important difference (MCID) in DAOH and its relation to clinically important long-term outcomes has not yet been studied for patients admitted to hospital for coronary artery bypass graft surgery (CABG), acute coronary syndrome (ACS), or heart failure (HF). We propose to determine whether differences in DAOH in common use as a MCID in clinical trials will be associated with significant differences in clinically important outcomes.

**Methods and Analysis:** This is a retrospective observational cohort study in three separate cohorts of adult (≥18 years) patients admitted to National Health Service (NHS) hospitals in England for: i) ACS ii) CABG iii) HF. Patients will be identified through Hospital Episode Statistics (HES) Admitted Patient Care (APC) data from 01/01/2009 – 31/12/2015 and followed up to 5 years after the index admission date.

Adjusted and unadjusted multivariable fractional polynomial Cox regression models will be used to estimate HRs for primary outcomes, according to pre-specified differences in DAOH.

**Ethics and Dissemination:** This is a sub study of the observational cohort study ‘In Silico Trials of Surgical Interventions - Using Routinely Collected Data to Model Trial Feasibility and Design Efficiency In Vivo Randomised Controlled Trials’ - ClinicalTrials.gov Identifier: NCT05853536. Ethical approval has been obtained from University of Leicester Research Ethics Committee (22322-yll15-ls:cardiovascularsciences). Findings from this study will be disseminated through peer-reviewed scientific publications and research conferences.

## Introduction

### Background and Rationale

**Outcomes** used in clinical trials traditionally measure important clinical events but may fail to capture outcomes more important to patients such as rehabilitation and healthcare related quality of life. There is increasing recognition that it is important to measure patient centred outcomes in a standardised and reproducible manner.

Systematic reviews of outcome measures used in cardiovascular research have identified that there is currently no accepted standard for reporting outcomes following a variety of interventions, such as after cardiac surgery.(1) This has resulted in heterogeneity in the outcome measures selected between trials in the same field, which reduces the ability to perform meta-analyses.(2) The COMET initiative supports the development of core outcome sets for trials in a specific clinical area. In a systematic review of systematic reviews of non-minimal-invasive cardiothoracic surgery RCTs, registered under the COMET initiative, Benstoem et. al. found that no patient centred outcomes were included in any meta-analysis and one third or reviews included no patient centred outcomes.(3) They conducted a Delphi consensus study to establish a core outcome set in pre-, intra- or postsurgical interventions in invasive off- or on-pump cardiac surgery. They recommend mortality, quality of life, hospitalisation and cerebrovascular complication as core outcomes to represent death, life impact, resource use and pathophysiological manifestations respectively.

Furthermore, most analyses use a time to first event as a composite endpoint. These endpoints have limitations, the major one being that the events within the composite measure have differing severities.(4, 5) Therefore, there is a research gap to develop patient-centred outcome measures and better standardise the analysis and reporting of outcome measures in cardiovascular clinical trials.

‘Days alive and out of hospital’ (DAOH) is a composite patient centred outcome measure that integrates several clinically important outcomes, including death, hospital length-of-stay, and hospital readmission.(6) It is easily quantifiable, incorporates re-admission and early deaths following discharge, and is popular with patients.

The “Standardised Endpoints in Perioperative Medicine initiative” recently recommended the use of DAOH at 30 days (DAOH_30_), following their systematic review and Delphi consensus process. DAOH_30_ was the only life impact measure recommended.(7)

DAOH has been validated as an outcome measure in some cohorts. Jerath et. al found a “poor” DAOH, defined as those with DAOH in the lowest tenth percentile, was found to be associated with a significantly increased incidence of post operative complications in their cohort study of non-cardiac surgical patients. The median DAOH_30_ in the lower tenth percentile was 16 days compared with a median of 26 days in the rest of the cohort.(6) Traditional risk factors for mortality and complications such as increasing age, worsening functional status, and increased duration of surgery have also been found to be significantly associated with lower median DAOH.(8, 9) In the pre-specified analysis of the ISCHEMIA trial, White et. al found there to be a 2.4 days difference in DAOH_30_ between invasive and conservative management in stable coronary disease with moderate or severe ischaemia, favouring conservative management.(10) However, there was no difference in the primary outcome; all-cause mortality in this trial. This creates uncertainty as to the clinical importance of differences in DAOH. For example, should differences in DAOH between treatment groups in randomised control trials (RCTs) lead to changes in practice in isolation?

We have reviewed all randomised control trials in acute coronary syndrome (ACS), heart failure (HF), and cardiac surgery patients with DAOH as an outcome measure up until 26/05/23. Studies were identified from MEDLINE database searches. From 106 RCTs where DAOH is used as an outcome, we identified 3 trials in ACS, 21 trials in HF and 4 trials in cardiac surgery. There was variation in measurement of DAOH, with 5 of the trials in HF not specifying the time of measurement.

Only 2 trials in heart failure cohorts reported a statistically significant difference in DAOH between treatment arms. In their analysis of the ESCAPE trial, Kalogeropoulos et. al. found a statistically significant reduction in median DAOH_180_ with in hospital inotrope use compared to controls (144 [IQR 135 - 167]) vs 165 [IQR 139 - 174]; P<0.001). This trial was powered to detect was an 8 day difference in DAOH_180_.(11)

Nieminen et. al. in their trial of levosimendan vs dobutamine for low output heart failure patients found a statistically significant 24-day improvement (157 vs 133 days, p = 0.027) in mean DAOH_180_. This study did not report a sample size calculation or specified MCID.(12) Three randomised control trials in HF with negative findings included DAOH as a primary endpoint and specified an MCID for a sample size calculation. The ELISABETH Randomised Clinical Trial, investigating the effect of an emergency care bundle among elderly patients with acute heart failure used DAOH_30_ as their primary endpoint. They calculated their sample size based on ≥ 3 days difference in DAOH_30_.(13)

The TELEREH-HF randomised control trial of hybrid comprehensive telerehabilitation vs usual care found a 1-day difference in median DAOH (775 vs 776) between treatment and control groups respectively. They used a 21-day difference in DAOH as their MCID for their sample size calculation over follow up from 12 to 24 months.(14)

The PRIMA study, investigated management guided by an individualised NT-proBNP target in patients hospitalised for decompensated, symptomatic HF with elevated NT-proBNP levels at admission used DAOH as their primary outcome over 2 years follow up. They found a non-statistically significant difference of 21 days in median DAOH (685 vs 664, p = 0.49) between treatment and control groups respectively. Their primary outcome was changed prior to recruitment and DAOH was not used in their power analysis. A post hoc analysis showed that a 4% difference in percentage of DAOH could be detected, which is approximately 28 days.(15)

There were two further randomised control trials in heart failure which included DAOH as the primary outcome but did not include a sample size estimate. They cited a lack of sufficient data in the literature to estimate an appropriate sample size.(16, 17)

Of trials relating to cardiac surgery, only 2 specify the time period for DAOH, both studies are trial protocols. The ITACS trial, conducted by Myles et. al. is powered to detect a median difference of 1.45 days for DAOH_90_ and 1.5 days for DAOH_30_.(18) The NOTACS trial, conducted by Earwaker et. al. is powered to detect a 2 day difference in DAOH_90_.(19)

There were 3 trials relating to ACS with DAOH as an outcome measure. The pre-specified analysis of the ISCHEMIA trial by White et. al. found a statistically significant 2.4 days difference in DAOH_30_ (28.4 vs 30.8, P < .001) and 6.3 days difference in mean DAOH_365_ (355.9 vs 362.2, P < .001) between patients treated with invasive or conservative management in stable coronary disease with moderate or severe ischaemia, favouring conservative management. In their analysis of TRILOGY ACS, which compared prasugrel versus clopidogrel in stable patients after non-ST segment elevation acute coronary syndrome treated without revascularization, Faranoff et. al. found a non-significant 1-day difference in mean DAOH_365_ (316 vs 317 p=0.67) between prasugrel and clopidogrel respectively. In their analysis of ODYSSEY OUTCOMES, which compared alirocumab with placebo when added to a high intensity statin after ACS, Szarek et. al. found 3-day difference in mean DAOH after ≥ 2 years follow up favouring alirocumab (1040 vs 1037, p = 0.05). None of these trials in ACS included DAOH as a primary outcome or specified a MCID for sample size calculation.(10, 20, 21)

In summary, the minimum clinical important difference (MCID) in DAOH and its relation to clinically important long-term outcomes has not been studied in ACS, CABG and HF patients. Understanding what differences in DAOH mean in terms of clinically important outcomes will enable better interpretation of clinical trials where this outcome measure is used, with regard to their likely impact on practice. This study will define the MCID in DAOH for these cohorts and allow for trials to be designed to detect a MCID that reflects clinically important outcomes.

## Objectives

Our primary hypothesis is that differences in DAOH in common use as MCID in clinical trials will be associated with significant differences in clinically important outcomes at later time points. For the purposes of this analysis, we hypothesise that:

1. A 2-day decrease in DAOH_90_ will be associated with a HR of >1.25 for the primary outcomes at 1 year in ACS and CABG patients.
2. A 4-day decrease in DAOH_90_ will be associated with a HR of >1.25 for the primary outcomes at 1 year in HF patients.
3. A 4-day decrease in DAOH_180_ will be associated with a HR of >1.25 the primary outcomes at 1 year in ACS and CABG patients.
4. An 8-day decrease in DAOH_180_ will be associated with a HR of >1.25 for the primary outcomes at 1 year in HF patients.
5. A 6-day decrease in DAOH_365_ will be associated with a HR of >1.25 for the primary outcomes at 5 years in ACS and CABG patients.
6. A 10-day decrease in DAOH_365_ will be associated with a HR of >1.25 for the primary outcomes at 5 years in HF patients.

Secondary hypotheses:

1. The decrease in DAOH specified for the primary analyses will be associated with statistically significant reductions in the individual components of the primary outcome.
2. There may be a non-linear relationship between DAOH and primary outcomes.

Our primary objectives are:

1. Calculate the DAOH_90_, DAOH_180_, DAOH_365_ in each cohort and time-to event estimates for specified primary outcomes.
2. Determine the time-to event estimates for primary outcomes for a: 2-day decrease in DAOH_90_, 4-day decrease in DAOH_180_ and 6-day decrease in DAOH_365_ in ACS and CABG cohorts.
3. Determine the time-to-event estimates for primary outcomes for a: 4-day decrease in DAOH_90_, 8-day decrease in DAOH_180_ and 10-day decrease in DAOH_365_ in a HF cohort.
4. Determine the time-to event estimate for secondary outcomes for a: 2-day decrease in DAOH_90_, 4-day decrease in DAOH_180_ and 6-day decrease in DAOH_365_ in ACS and CABG cohorts.
5. Determine the time-to-event estimates for secondary outcomes for a: 4-day decrease in DAOH_90_, 8-day decrease in DAOH_180_ and 10-day decrease in DAOH_365_ in a HF cohort.
6. Explore the (non-linear) association between differences in DAOH as a continuous measure and the time-to-event of primary outcomes.

## Methods

### Design, Setting, Participants

This will be a retrospective observational cohort study in three separate cohorts of adult (≥18 years) patients admitted to National Health Service (NHS) hospitals in England for: i) Acute coronary syndrome (ACS) ii) Coronary artery bypass grafting (CABG) iii) Heart failure (HF). Patients with an ICD-10 or OPCS4 code indicating ACS, CABG procedure and HF between 01/01/2009 – 31/12/2015 will be identified and followed up to 5 years after the index admission date. Information on admissions 2 years prior to the index date will also be collected for ascertainment of baseline demographics and comorbid conditions.

### Steps to Mitigate Bias

The use of national datasets with high levels (>96%) of accuracy for procedures and diagnostic codes will reduce selection and detection bias.(22, 23) This protocol will be published prospectively in a publicly available registry to prevent reporting bias. The analysis will be reported according to the Strengthening the Reporting of Observational Studies in Epidemiology (STROBE) initiative reporting standards.(24)

Potential confounding variables on outcomes which are available in Hospital Episode Statistics (HES) data (age, sex, ethnicity, socioeconomic deprivation, prior myocardial infarction (MI), prior diabetes, prior hypertension, prior lipidaemia, prior cerebrovascular disease, prior chronic kidney disease, Charlson comorbidity index, admission method) will be adjusted for in our statistical models.

### Data Sources

Our cohorts will be created using data from:

i. HES Admitted Patient Care (APC) records for hospitals in England 2007-2020.
ii. Office for National Statistics Civil Registrations data 2009 – 2020.

The HES database contains administrative data from English hospitals in the NHS. This includes records of inpatient admissions, outpatient appointments and accident and emergency (A&E) attendances. HES APC records include information on episodes of treatment that require a hospital bed. These could be emergency or elective admissions.(25)

HES APC records will be matched with the ONS civil registrations data set for mortality outcomes by anonymised unique patient ID. HES APC records contain information on diagnoses, procedures, age, sex, ethnicity, socioeconomic deprivation, index hospital admission, admission method, discharge dates, re-admission (and subsequent discharge) dates.

HES Critical Care (CC) records will not be included in the creation of our cohorts and therefore details of critical care or intensive care inpatient stays will not included in this analysis.

The requested HES datasets has been pseudonymised with unique identifiers generated by NHS Digital. Death and DID datasets are linked to the HES datasets through bridging files provided by NHS Digital. In addition, patient records will be longitudinally linked through the pseudo identifiers.

### Data Considerations

Information on age, sex, ethnicity, socioeconomic deprivation (measured by Index of Multiple Deprivation), and admission method from the index episode will be used. The Index of Multiple Deprivation is made up from 7 domains - income deprivation, employment deprivation, health deprivation and disability, education skills and training deprivation, barriers to housing and services, living environment deprivation, and crime.(26) Patients will be grouped into one of ten deciles of IMD from the most to least deprived. Ethnicity will be categorised as White, Black, Asian, Mixed/Other, Unknown.

Prior comorbidities (prior MI, diabetes, hypertension, lipidaemia, cerebrovascular disease, chronic kidney disease) will be identified from the presence of at least one characteristic ICD-10 or OPCS4 code from admissions 2 years prior to the index date.(27) Additional comorbidities will also be identified, to calculate the Charlson comorbidity index. These include: peripheral vascular disease, congestive heart failure, dementia, chronic pulmonary disease, rheumatologic disease, hemiplegia or paraplegia, liver disease, other renal disease, tumour, leukaemia or lymphoma, metastatic solid tumour, HIV or AIDS.

The ICD-10 and OPCS-4 codes are in Appendix 1.

### Exposures

The primary exposures will be DAOH at; 90 days (DAOH_90_), 180 days (DAOH_180_), and 1 year (DAOH_365_).

DOAH will be defined as the number of days alive and out of hospital within a defined period, starting at the day of admission (day 0). DAOH will be calculated in the following way: (Number of days in the study period) – (hospital days + mortality days). The defined number of days for each study period is 90 days, 180 days and 365 days for DAOH_90_, DAOH_180_, DAOH_365_ respectively. Hospital days is defined as the number of days spent in the inpatient setting, identified through HES APC records. Mortality days is defined as “the number of days remaining in the observation period following the date of death”. For example, for DAOH_365_, a patient who was alive 1 year after the admission date would have zero mortality days while a patient who died 100 days after the admission date would have 265 mortality days.(28)

In sensitivity analyses we will also report an alternative calculation of DAOH described by Myles et. al. where patients who have died before the end of the study period are assigned DAOH = 0 for that respective period.(8) We will compare this with our primary analysis findings.

### Outcome Variables

#### Primary Outcomes

For the ACS cohort, the primary outcome for DAOH_90_ and DAOH_180_ will be major adverse cardiac events (death, stroke, MI or repeat revascularisation; MACE) at 1 year. The primary outcome for DAOH_365_ in the ACS cohort will be MACE at 5 years.

For the CABG cohort, the primary outcome for DAOH_90_ and DAOH_180_ will be MACE at 1 year. The primary outcome for DAOH_365_ in the CABG cohort will be MACE at 5 years.

For the HF cohort, the primary outcome for DAOH_90_ and DAOH_180_ will be a composite of cardiovascular re-hospitalisation or all-cause death at 1 year. The primary outcome for DAOH_365_ in the HF cohort will be a composite of cardiovascular re-hospitalisation or all-cause death or at 5 years.

#### Secondary outcomes will be

1. Myocardial infarction *at 1 and 5 years*.
2. Stroke *at 1 and 5 years*.
3. Revascularisation *at 1 and 5 years*.
4. All cause death *at 1 and 5 years*.
5. Cardiovascular rehospitalisation *at 1 and 5 years*.

5-year outcomes will only be assessed for DAOH_365_.

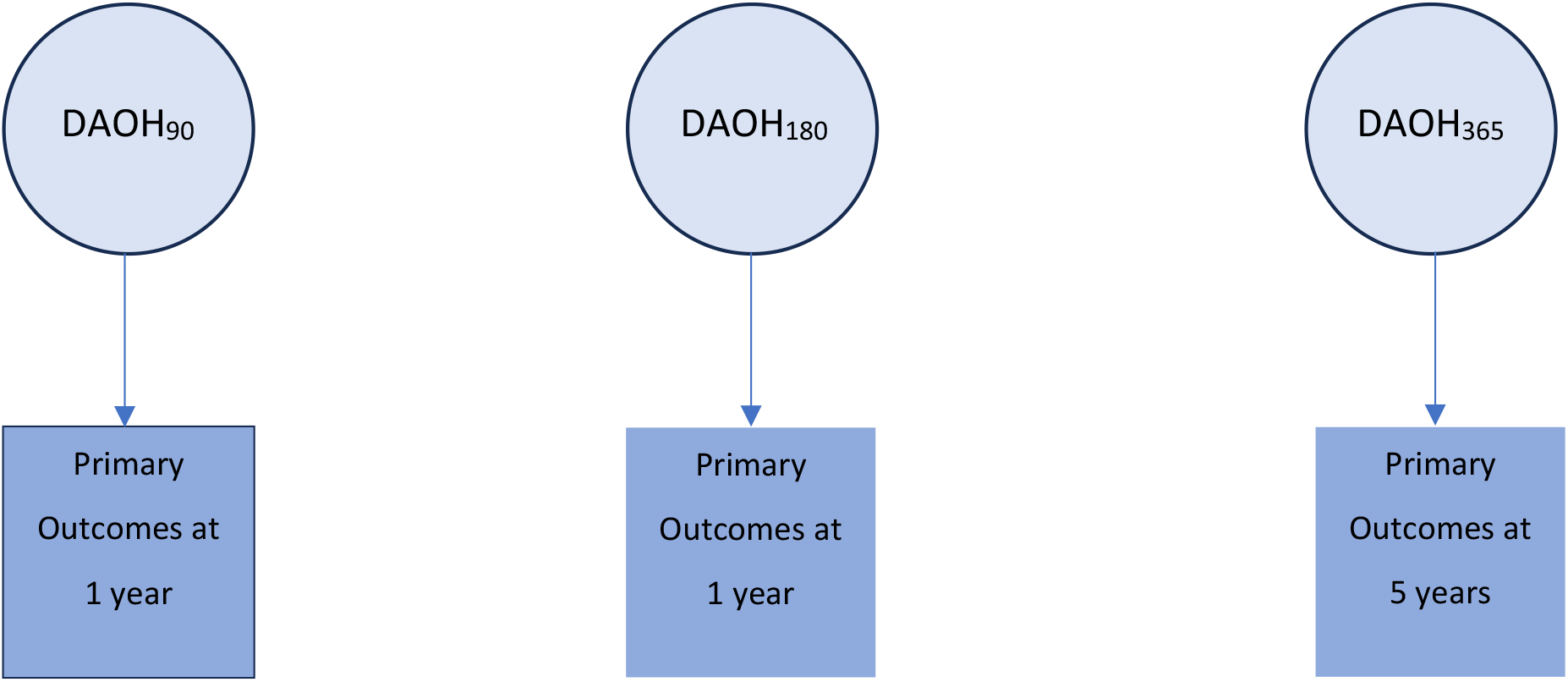

The date and cause of death will be identified from ONS mortality data, linked to HES admissions data. All-cause mortality and cardiovascular mortality will be identified at 1 year and 5 years from the index admission date. Cardiovascular hospitalisation and readmission for heart failure, MI, stroke, revascularisation (defined as a composite of CABG and PCI) and MACE (defined as a composite of MI, stroke, revascularisation and cardiovascular death) will be identified at 1 year and 5 years from the index date.

Survival time will be calculated from the index admission date to occurrence of the first primary event. Censoring will be applied to event free patients at the end of the follow up period or those who have died from an unrelated event.

## Statistical Methods

### Primary Analysis

1. We will calculate DAOH_90_, DAOH_180_, DAOH_365_, in each cohort, and present as a histogram with median (Q1, Q3), mean and SD.
2. We will report death rates per 1000 people, index admission hospital length of stay (mean) and number of re-admissions (mean) in each cohort.
3. We will produce Kaplan Meier survival curves for the primary and secondary outcomes in all three patient cohorts.
4. We will fit a multivariable fractional polynomial Cox regression model for primary outcomes in each cohort for our pre-specified differences in DAOH. We will adjust for: age, sex, ethnicity, index of multiple deprivation, admission type, comorbidities and Charlson comorbidity index.
5. To assess the HR for a 2-days difference in ACS and CABG cohorts, we will create three DAOH_90_ categories: i) median DAOH_90_ ± 1 day ii) median DAOH_90_ + 1 day iii) median DAOH_90_ - 1 day. We will calculate the adjusted HR for each category. We will replicate this analysis for a 4-day difference in DAOH_180_ and 6-day difference in DAOH_365_ for ACS and CABG cohorts. We will replicate this analysis for a 4-day difference in DAOH_90_, 8-day difference in DAOH_180_ and 10-day difference in DAOH_365_ for the HF cohort.

For the purposes of the analyses, an acceptable DAOH MCID will be defined as a that which gives a statistically significant HR ≤0.8 or ≥1.25 (1/0.8) for the primary outcome.

### Secondary Analysis

Cox proportional hazards models will be fitted for the primary outcome stratified by DAOH. We will choose the DAOH strata cut points based on the distribution of DAOH_90_, DAOH_180_ and DAOH_365_ in each cohort. We will select the strata with the highest DAOH as the reference group for our analysis and calculate unadjusted and adjusted hazards ratios for each group, adjusting for: age, sex, ethnicity, index of multiple deprivation, admission type, comorbidities and Charlson comorbidity index.(29)

### Subgroup Analysis

To explore heterogeneity of effect of DAOH on primary outcomes, we will conduct a subgroup analysis. The pre-specified patient subgroups will be: urgent surgery, elective surgery (CABG cohort), ST-elevation myocardial infarction (STEMI), Non-ST elevation myocardial infarction (NSTEMI) (ACS cohort), patients with multiple (2+) long-term conditions and ages <65, 65-75 and >75 (all cohorts). These subgroups have been chosen based on clinical reasoning, as they are likely to have different risk of mortality or adverse outcomes. Expected high risk groups would be urgent surgery, STEMI, patients with multiple long-term conditions and ages >75.

### Sensitivity Analysis

An alternative calculation of DAOH, where patients who have died before the end of the study period are assigned DAOH = 0 has been previously published. We will also calculate the DAOH_90_, DAOH_180_, DAOH_365_, in each cohort using the alternative method, described by Myles et. al., where death during the study period is assigned a DAOH = 0. We will present this as a histogram with median (Q1, Q3), mean and SD.(8)

As a sensitivity analysis we will repeat our modelling of the primary outcome using this method in each cohort.

### Exploratory Analysis

We will explore the relationship between numeric DAOH_90_, DAOH_180_, and DAOH_365_ with their respective primary outcomes for each cohort, which could be non-linear. We will use a multivariable fractional polynomial Cox regression model for the hazard of primary outcomes. We will adjust for: age, sex, ethnicity, index of multiple deprivation, admission type, comorbidities and Charlson comorbidity index. Statistical analysis will be conducted using R-Studio (version 4.2.3) and Stata 18.0.

## Data Handling and Record Keeping

### Approvals and Data Handling

Access to the data will be restricted to delegated researchers and statisticians in this project who are staff or Ph.D. students of the University of Leicester. The Chief Investigator will be responsible for applying and maintaining appropriate access rights to the datasets. Staff will adhere to a Data Management Protocol which outlines responsibilities and training requirements.

The University of Leicester has a current NHS Digital Data Sharing Framework Contract (CON-313050-C7D3F) and will act as the Data Controller for the NHS Digital datasets relating to this project. The data protection registration number for the University of Leicester is Z6551415. The University Information Security Policy is available at: https://uniofleicester.sharepoint.com/sites/staff/information-assurance-services/SitePages/Home.aspx

The University of Leicester College of Life Sciences has submitted the first NHS Digital Data Security & Protection Toolkit in March 2019 (code EE133832-CMBSP), and annually thereafter.

### Dataset Creation

Each cohort will be created using HES APC records. We will identify patients and records of interest based on ICD-10 and OPCS4 codes within the dataset and merge across years to create out cohort for analysis. We will use HES APC records at the time of admission and from 2 years prior to the index date to create our variables for analysis. We will use records from the admission date to calculate DAOH. We will merge with ONS mortality data using the non-identifiable “TOKEN_PERSON_ID” to identify patient deaths in our follow up period.

Data extraction will be done by Robert Grant (Clinical Research Fellow).

### Data Storage

All data and documentation related to the project is stored on the secure dedicated research data storage service known as the Research File Store (RFS) at the University of Leicester. The RFS is a secure and resilient server that adheres to current information governance standards and is centrally managed by the University of Leicester to ensure it is updated to meet future changes in data security standards. The RFS is based on enterprise class storage. There are no removable media or systems in the solution. The RFS is housed in two secure data centres which are access controlled via swipe card and pin and monitored via CCTV. Access is restricted to essential IT Services staff. Any third-party access is supervised. The RFS is backed-up nightly to an enterprise-class backup facility in a further secure, access-controlled data centre. Backups are retained for a year in line with a Backup Retention schedule.

No copies of the data will be made to other locations. The University of Leicester holds Cyber Essentials certification for the RFS which is accessed from its fully managed desktop/laptop service. The University of Leicester Cyber Essentials certification number is QGCE597 and can be validated on the National Register of Cyber Essentials Certified Companies.

Remote work on the data will be via access to RFS using fully assured IT devices, via VPN.

### Data Processing

Data processing will be undertaken by delegated research fellows or PhD students and validated by the study statistician by the methods described under statistical methods.

### Data Destruction

Data retention and destruction will be subject to the active Data Sharing Agreement with NHS Digital. At end of life, all RFS servers, storage systems and desktop PCs are disposed of under the University Estates Division’s managed waste disposal contract to ensure the University’s compliance with its WEEE obligations. This contract engages a third-party organisation to securely wipe all disks. The contracted company uses specialised software to provide secure data destruction to U.S. DoD 5220.22-M, U.S. DoD 5220.25, U.S. DoD 5200.28M and HMG (CESG) IS5 baseline and enhanced.

### Access to Data

Direct access will be granted to authorised representatives from the Sponsor, host institution and the regulatory authorities to permit project-related monitoring, audits and inspections.

At least annually there is an audit of R Drive user accounts to confirm/alter users and permissions.

### Archiving

The final dataset for each cohort and analysis will be stored on the Research File Store.

## Monitoring, Audit & Inspection

The study will be conducted and monitored in accordance with the current approved protocol, ICH GCP, relevant regulations and standard operating procedures.

## Ethical and Regulatory Considerations

### Participant Confidentiality

Data from NHS Digital will be pseudonymised record-level data, and will not contain any identifiable data. The University will have no access to the file that can link the pseudo identifiers to the patients. As such the data is effectively anonymous.

### Project File

All correspondence regarding the project will be retained in the “In silico trials of surgical interventions - using routinely collected data to model trial feasibility and design efficiency in vivo RCTs” project file to allow reconstruction of the project in the future.

### Sponsor Standard Operating Procedures

All relevant Sponsor SOPs will be followed to ensure that this study complies with all relevant legislation and guidelines

### Declaration of Helsinki

The Investigator will ensure that this study is conducted in full conformity with the current revision of the Declaration of Helsinki (last amended October 2000, with additional footnotes added 2002 and 2004).

### ICH Guidelines for Good Clinical Practice

The Investigator will ensure that this study is conducted in full conformity with relevant regulations and with the ICH Guidelines for Good Clinical Practice (CPMP/ICH/135/95) July 1996.

### Annual Progress Reports

An annual progress report (APR) will be submitted to the Sponsor within 30 days of the anniversary date on which the ethics approval was given, and annually until the project is declared ended, and annual confirmation reports (ACR) to NHS Digital.

### Peer review

This protocol was reviewed and developed by the CI, delegated researchers and a study statistician.

### Protocol compliance and Serious Breaches

Prospective, planned deviations or waivers to the protocol are not allowed and must not be used. Accidental protocol deviations can happen at any time. They must be adequately documented on the relevant forms and reported to the Chief Investigator and Sponsor immediately. Deviations from the protocol which are found to frequently recur are not acceptable, will require immediate action and could potentially be classified as a serious breach.

### Indemnity

University of Leicester’s insurance applies.

### Access to the final project dataset

Access to the final project dataset will be granted to the CI, delegated researchers, and the study statistician.

### Dissemination

All analyses are conducted on de-identified data. We will only present summary statistics with no individual level of information.

Scientific findings will be disseminated by usual academic channels, i.e. presentation at international meetings and by peer-reviewed publications, where available.

## Data Availability

The data underlying this article were provided by NHS Digital under licence. The data that support the findings of this study are available from the corresponding author upon reasonable request. Raw data may be shared with permission of NHS Digital.

