## Appendix 1 for "Validation of “Days Alive and out of Hospital” as an Outcome Measure After Coronary Artery Bypass Graft Surgery, Acute Coronary Syndrome and Heart Failure Hospitalisation"

**OPCS and ICD codes for Phenotyping:**

Acute coronary syndrome will be defined by ICD codes: I20.0, I21, I22, I24.(1)

STEMI will be defined by ICD codes: I21.0 - I21.3, I22.0, I22.1, I22.8, I22.9.(2)

NSTEMI will be defined by ICD codes: I20.0, I21.4, I22.2.(2)

Coronary artery bypass grafting will be defined by OPCS 4.10 codes: K40 – 46.(3)

Heart failure will be defined by ICD code I50.(1)

Major adverse cardiac events will be defined as: acute myocardial infarction (ICD I21, I22), acute stroke (ICD I61 – I64), cardiovascular death (ONS cause of death ICD I00 to I99) and revascularisation (OPCS4 K40 - 46, K49 – 50, K75).

Revascularisation will be defined by the following OPCS4 codes: K40 – 46, K49 – 50, K75.(4)

Cardiovascular hospitalisation will be defined as all admissions with cardiovascular causes as the primary diagnostic code (ICD I00 to I99).(3)

Heart failure hospitalisation will be defined by ICD codes: I11.0, I13.0, I13.2, I126.0, I15.0.(3)

Diabetes will be defined by ICD codes: E10 – E14.

Hypertension will be defined by ICD codes I10 – I13,  I15.

Chronic kidney disease will be defined by N18, N19, N990, N03, I13.

Lipidaemia will be defined by ICD codes E78.(3)

ICD codes for the Charlson Comorbidity Index: myocardial infarction (I21, I22, I25.2), congestive heart failure (I09.9, I11.0, I13.0, I13.2, I25.5, I42.0, I42.5 – I42.9, I43, I50, P29.0), peripheral vascular disease (I70, I71, I73.1, I73.8, I73.9, I77.1, I79.0, I79.2, K55.1, K55.8, K55.9, Z95.8, Z95.9), cerebrovascular disease (G45, G46, I60 – I69, H34.0), dementia (F00 – F03, G30, F05.1, G31.1), chronic pulmonary disease (I27.8, I27.9, J40 – J47, J60 – J67, J68.4, J70.1, J70.3), rheumatologic disease (M05, M06, M31.5, M32 – M34, M35.1, M35.3, M36.0), peptic ulcer (K25 – K28), hemiplegia or paraplegia (G04.1, G11.4, G80.1, G80.2, G81, G82, G83.0, G83.1, G83.2, G83.3, G83.4, G83.9), diabetes without complications (E10.0, E10.1, E10.6, E10.8, E10.9, E11.0, E11.1, E11.6, E11.8, E11.9, E12.0, E12.1, E12.6, E12.8, E12.9,

E13.0, E13.1, E13.6, E13.8, E13.9, E14.0, E14.1, E14.6, E14.8, E14.9), diabetes with chronic complications (E10.2 – E10.5, E10.7, E11.2, E11.5, E11.7, E12.2 – E12.5, E12.7, E13.2 – E13.5, E13.7, E14.2 – E14.5, E14.7), mild liver disease (B18, K70.0 – K70.3, K70.9, K71.3 – K71.5, K71.7, K7.3, K7.4, K76.0, K76.2 – K76.4, K76.8, K76.9, Z94.4), moderate to severe liver disease (I85.0, I85.9, I86.4, I98.2, K70.4, K71.1, K72.1, K72.9, K76.5 – K76.7), renal disease (I12.0, I13.1, N03.2 -N03.7, N05.2 – N05.7, N18, N19, N25.0, Z49.0 – Z49.2, Z94.0, Z99.2), tumour, leukaemia or lymphoma (C00 – C26, C30 – C34, C37 – C41, C43, C45 – C58, C60 – C76, C81 – C85, C88, C90 – C97), metastatic solid tumour (C77 – C80), HIV or AIDS (B20 – B22, B24).(5)

OPCS4 codes for the Charlson comorbidity index: cerebrovascular disease (L29, L31.1, L31.2), peripheral vascular disease (L16, L20-21, L23, L25, L27.1-3, L27.6-9, L51-52, L54, L59-60, L634, X07-11).(6)
